# Automated Data Extraction Model for the USIDNET Registry: Bigger, Faster, and Better Data Collection

**DOI:** 10.1101/2025.08.25.25333562

**Authors:** Vaibhavi Vichare, Gonench Kilich, Charlotte Cunningham-Rundles, Michelle Onyekaba, Roshini S. Abraham, Jennifer Puck, Ramsay Fuleihan, Rebecca Marsh, Kathleen E Sullivan

## Abstract

The United States Immunodeficiency Network (USIDNET) is an NIH-funded research consortium that advances scientific investigation on inborn errors of immunity (IEI). Formerly, USIDNET Registry data were collected via an opt-in system with patient informed consent. The current Registry uses semi-automated, de-identified data extraction from EPIC with a consent waiver. This study was performed to assess whether the new method improved enrollment using data from one site. Diagnoses, sex, and age within the new (n= 1145) and old (n= 551) registries were defined. The new registry enrolled twice and many subjects and had six times more clinical features recorded per patient on average and 22 times more laboratory data recorded per patient. Response to queries is much more rapid, with execution of a database query within a day. There were differences in enrollment demographics depending on underlying diagnosis. The design of the new USIDNET Registry may better capture a greater number and representation of patients compared to the old Registry.

**Summary:** USIDNET is a suite of resources for clinical immunologists. The Registry of patient data utilizes data extraction from electronic health records to minimize burden on participating sites. This has been effective at improving enrollment.

## Introduction

The United States Immunodeficiency Network (USIDNET) was designed to provide multiple resources to the Clinical Immunology community. A Registry is a central component of the suite of resources. The previous version of the USIDNET Registry was comprised of over 5000 IEI patients from 43 centers (1). That Registry collected clinical, genetic, and phenotypic patient data via a clinician accessed RedCap data entry system. USIDNET Registry data has been used as a scientific resource for critical research on the molecular and genetic pathways involved in IEI biology (selected examples (2-14), however, longitudinal data were unstructured and incompletely provided. Antibody deficiencies represented about half of all patients in the previous Registry. A goal of USIDENT has been to improve longitudinal data collection, data completeness, and patient representation. Longitudinal data collection will improve outcomes research and increasing enrollment and types of patients is critical to define IEI phenotypes among different populations. A 2024 study by O’Connell and colleagues compared a cohort of pediatric patients diagnosed with IEI to a comparator congenital heart disease (CHD) cohort and found that while the CHD cohort demographics mirrored the local population, the IEI cohort was disproportionately comprised of European ancestry patients with private insurance. Specifically, they found varying diagnostic rates of IEI between different demographics and differences associated with emergency room visits and hospitalizations (15). Another study found deaths related to IEI had the highest rate for non-European ancestry patients (16). Challenges related to obtaining a referral to a tertiary care specialist to receive an IEI diagnosis may contribute to some differences in outcomes among health disparity populations (17). Recognizing these challenges, we set out to try to improve data collection such that USIDNET would improve enrollment.

The new USIDNET Registry uses automated data extraction to improve longitudinal data and a waiver of consent to improve representation of the clinic population at participating sites. This study was performed at a single site since the current participating sites do not match the sites participating in the previous Registry. We compared diagnoses and enrollment and then compared to inpatient and outpatient populations as well as comparing the site’s demographics in the previous and current registries.

## Results

A goal of the current USIDNET Registry is to increase enrollment. In this study, we hypothesized that implementing a semi-automated data extraction method from the electronic medical record with a waiver of consent would improve enrollment. The new Registry roughly doubled enrolment at this one site from 551 in 2019 to 1145 enrollees in 2025. This represents an 82% capture of current patients identified as immunodeficient at this institution. We assessed disease categories by physician definition in the previous USIDNET Registry and ICD code definition in the current Registry (Figure 1). The distribution was different. Overall, there were more antibody deficiency diagnoses with greater granularity in the new Registry. The density of data per patient also increased. As an example, the previous Registry had 131 unique patients with CVID. The average number of non-laboratory data points per patient was 13 compared to 60 per patient in the new registry which had 303 patients with CVID. Overall, the patients in the previous registry had 6.5 clinical features recorded per patient and 26 lab results per patient. In the new registry, the patients had 38.5 clinical features and 866 lab values on average (p<0.0001 for both). This represents a six-fold increase in clinical features recorded per patient and a 22-fold increase in laboratory data.

**Figure 1.**
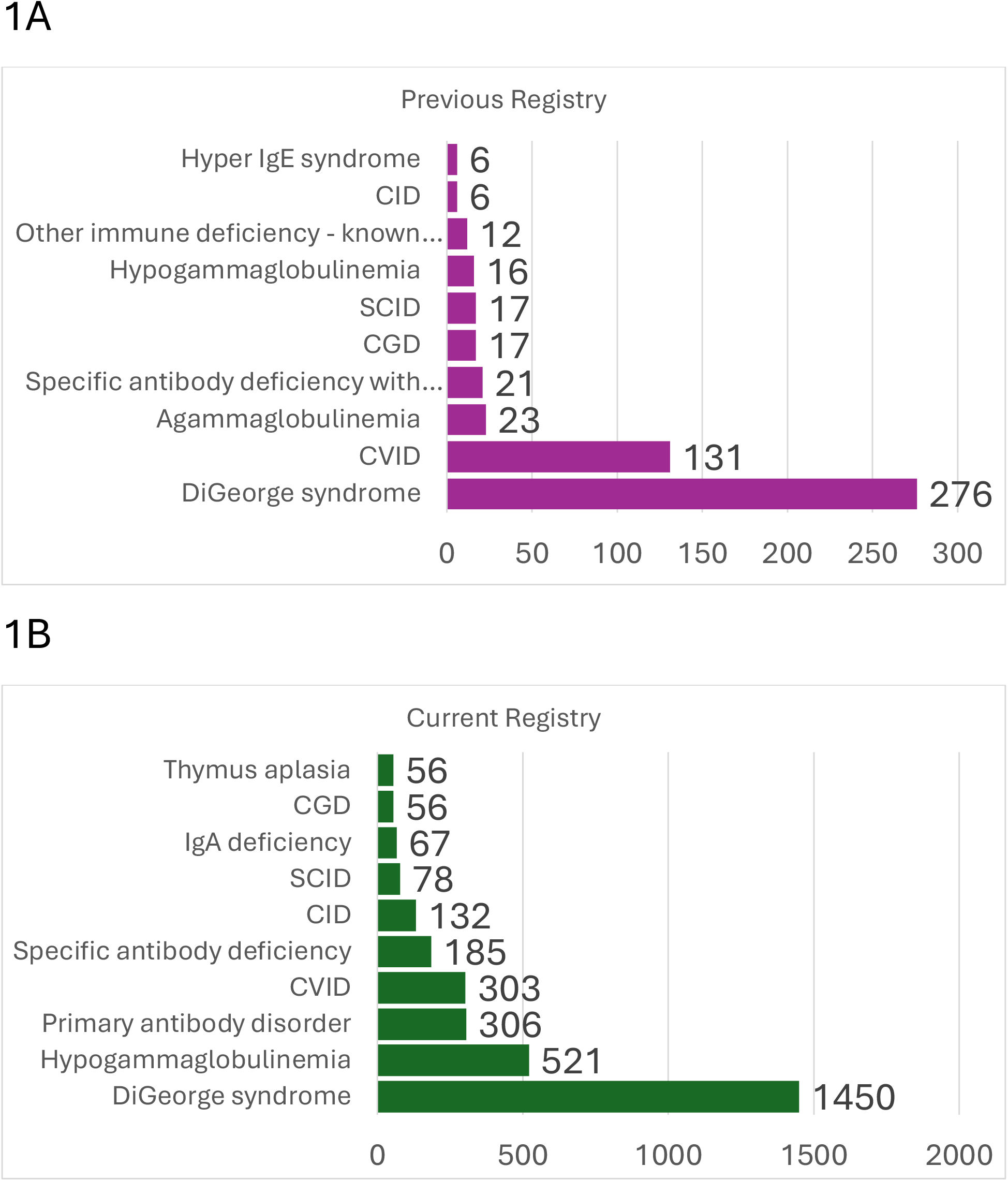
The ten most common diagnoses in each Registry. These two graphs list unique patient counts. A) The most common diagnoses in the previous Registry were DiGeorge Syndrome (n= 276), Common Variable Immunodeficiency (CVID) (n= 131), Agammaglobulinemia (n= 23), Severe Combined Immunodeficiency (n= 17), and Chronic Granulomatous Disease (n=17). **B)** The most common diagnoses in the current Registry (n= 2796) were DiGeorge Syndrome (N=1450), Hypogammaglobulinemia (N=521), Primary Antibody Disorder (N=306), Common Variable Immunodeficiency (CVID) (N=303), and Specific Antibody Deficiency (N=185).

The structure of the previous Registry required significant manual data curation of the output. The data queries can be completed in less than a day from the new Registry.

We additionally hypothesized that the demographics of the enrolled patients would change under the new model. The median age in the previous Registry was 28 years of age (minimum 9 years, maximum 72 years of age) at time the Registry closed to enrollment (2020). The median age in the current Registry was 17 years of age (minimum 0 years, maximum 54 years of age) (data not shown). The previous Registry was 63.7% male and the current Registry was 61.6% male. We found that there was a statistically significant (p <0.0001) change in the distribution of different populations in the new cohort as compared to the old, with an increase in the proportion of people of color with the largest increase in people identifying as “More than one race” (Figure 2A). The population of people of Latin ancestry changed very little between the two registries (Figure 2B).

**Figure 2.**
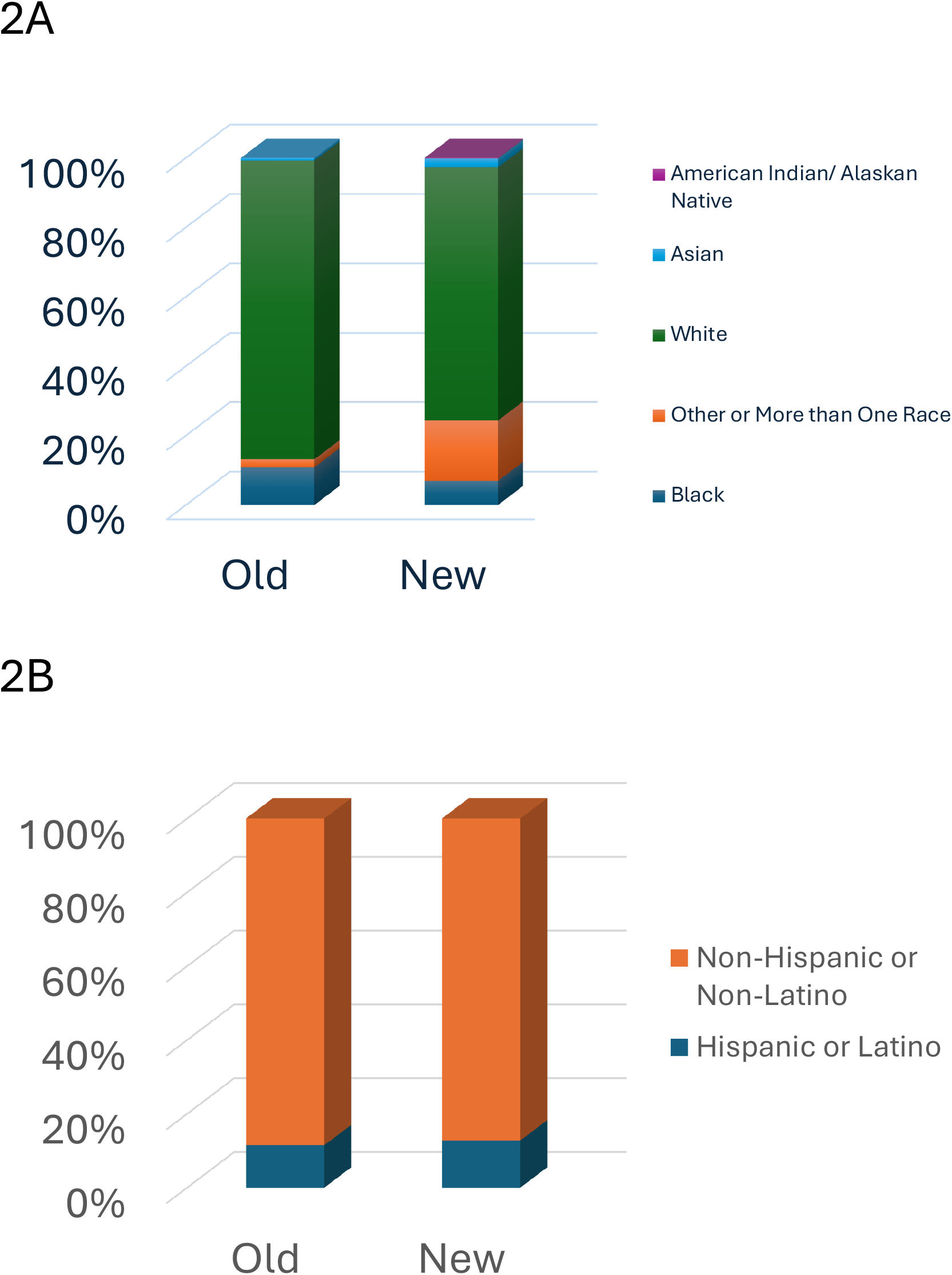
Demographics in previous and current Registries. **A)** The current (New) Registry data from the study institution includes 1138 subjects with racial information: 0.18% American Indian /Alaska Native, 2.55% Asian, 6.94% Black, 72.93% White, and 17.40% other or more than one race. The previous (Old) Registry includes 129 subjects: 0.78% Asian, 10.85% Black, 86.05% White, and 2.33% other or more than one race (Chi Square p< 0.0001). **B)** The current (New) Registry includes 1025 subjects with ethnicity information: 12.78% Hispanic/Latino, 87.22% Non-Hispanic/Latino. The previous (Old) Registry included 113 subjects with ethnicity information: 11.50% Hispanic/Latino, 88.50% Non-Hispanic/Latino (Chi Square p = 0.697)

To understand if there were disease-specific effects, we assessed the racial and ethnic representation in agammaglobulinemia, severe combined immunodeficiency (SCID), and chronic granulomatous disease (CGD) in both cohorts. In the current Registry, all patients with SCID would have been detected through newborn screening and therefore ascertainment should reflect the population of the region. Agammaglobulinemia and CGD both require recognition and referral to a medical center, a process where challenges might intervene and lead to imperfect ascertainment. We found there was a statistically significant difference in distribution according to ancestry between the two registries for agammaglobulinemia (p=0.0194), but not for SCID (p=0.43) or CGD (p=0.62). The distribution in the three disease states was similar to the overall cohort distribution in the recent Registry with higher numbers of “More than one race” in all three disease categories than in the previous Registry. Latin heritage and sex distribution remained relatively unchanged (not shown).

To better understand the demographics of the broader population being treated within this institution, we looked at the breakdown of total visits between 2010 and 2024 at this one institution. We found that there were fewer patients of color seen in the Immunology, Gastroenterology, and Oncology sub-specialties relative to Neonatology and the Emergency Department (Figure 3). The current Registry demographics differed from the total population (p<0.0001) but not from the Immunology Clinic demographics. The total Immunology clinic visits included: 16.49% Black, 14.77% Other, 62.86% White, 3.36% Asian, 0.16% American Indian/Alaskan Native, and 1.46% Unknown.

**Figure 3.**
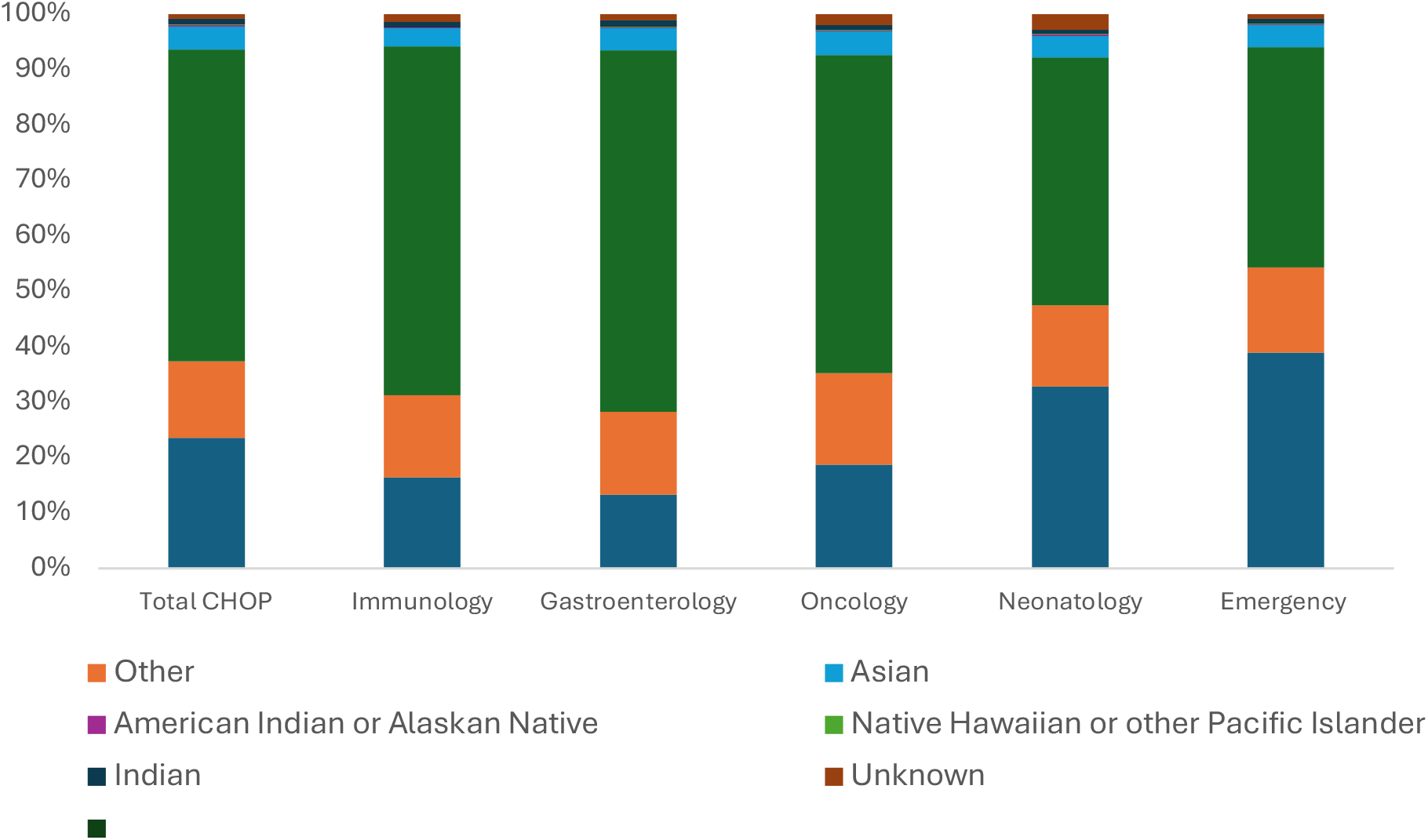
Comparison of demographics across diEerent clinical settings. CHOP visits were tabulated by department between 2010 and 2024. The demogaphic distribution varied across clinical sites with Neonatology and the Emergency Department having the highest rates of non-European ancestry patients. The difference between total CHOP and Immunology was significantly different with p< 0.0001, whereas the difference between Immunology and the current Registry was not significant.

## Discussion

We hypothesized that automated data extraction and a waiver of consent would remove some challenges to participation and improve the range of patients included in the USIDNET Registry. We noted that the top ten IEI diagnoses were different in the current Registry compared to the previous Registry. In the previous Registry, diagnosis entry was restricted by a pull-down menu that limited options, whereas the current Registry records all diagnoses available within the ICD code set. Thus, the more granular listing of antibody disorders likely reflects a methodologic change in data collection and suggests that USIDNET successfully expanded the range of diagnoses in the current version compared to the previous version.

Two clear improvements in expanding the range of patients were observed. The number of patients from this single institution doubled between the previous and current registries and the current registry collected 82% of all patients identified through ICD codes as having an immunodeficiency. It is not possible to define the variables governing this increase, but removing the burden of data entry through the use of an automated data extraction from the electronic medical record likely played a significant role as well as the necessity of obtaining consent.

The percentage of patients from non-European ancestry backgrounds also changed in the current Registry. It is useful to compare the demographics to other large populations in the US. In a study of national insurance claims, 83% of individuals were European ancestry (18). In a Chicago study using ICD codes, 63% of individuals with IEI were European ancestry (19) and a New York study using ICD codes found that 77% were European ancestry (20). The previous USIDNET Registry had 86% European ancestry participants with the current Registry having 73%.

The current Registry collects data back to 2018, a time frame when newborn screening for SCID was occurring in the state of Pennsylvania. Therefore, comprehensive capture of all patients in our catchment area would be expected. In the previous Registry, 13% non-European ancestry patients with SCID were recorded compared to 42% in the current Registry. This is similar to the overall clinic population of 37% non-European ancestry patients but somewhat less than the overall CHOP population with 47% non-European ancestry patients. For agammaglobulinemia, the previous Registry had 18% non-European ancestry patients compared to 57% in the current Registry. Thus, progress has been made in improving USIDNET enrollment at one institution.

These data demonstrate that the current Registry enrollment demographics are similar to the subspecialty clinics at this institution. However, we note a substantial difference between the three outpatient specialties analyzed compared to the total institutional population, the inpatient Neonatology population, and the Emergency Department population. There are likely multifactorial reasons for the USIDNET enrollees not matching the overall institutional demographics. Health disparity populations have less access to tertiary care, likely leading to fewer diagnoses of immunodeficiencies. We noted that Gastroenterology and Oncology had similar demographics, supporting an element of diminished access in general to subspecialists for people non-European ancestry. Ultimately, the new system of USIDNET data extraction led to a statistically significant increase in the enrollment of patients and improved data density.

This study examined an important question for all registries. Are the data representative of what clinicians will encounter in their practice and is it therefore of value? USIDNET has made major strides in improving number of patients, and demographic matching to the clinical anchor population, at least at one institution. Furthermore, data extraction is simpler and the longitudinal data are structured. This study does have some limitations, however. The enrollment in the previous Registry was smaller than the current Registry leading to small numbers of evaluable patients for some analytic categories. Nevertheless, these data support our hypothesis that representation will improve if challenges and hurdles are minimized for inclusion. There remains significant work to provide a fully representational data set, but the recent progress demonstrates that USIDNET provides data on a population of value to practicing clinicians.

## Methods

USIDNET leverages the Translational Data Warehouse (TDW) built on PostgreSQL. This infrastructure supports automated data extraction through standardized query templates applied to EPIC Clarity, allowing consistent and reliable integration of clinical data from participating sites. It is a standardize database with 15 core clinical concepts and captures more than 50 data fields including lab data, imaging, pathology, medications, procedures, and visit diagnoses. Demographic data is extracted directly from Clarity.

We analyzed data from a single site, Children’s Hospital of Philadelphia, that had participated in both registries and for which we could obtain demographic data across different clinical settings. For the previous USIDNET Children’s Hospital of Philadelphia Registry (n= 551), patient demographic data was obtained following a consent process and data entry by a trained research coordinator or a clinician. For the new Registry (n =1145), patient data was collected from the electronic health record (EHR) using a semi-automated data extraction model with a consent waiver. Patients without data for a given demographic were excluded from the analysis for that demographic. Demographic data on total CHOP patient visits and visits to immunology, gastroenterology, oncology, neonatology, and emergency medicine between 2010 and 2024 were ascertained using the SlicerDicer Epic tool. Chi-squared and Fisher’s exact test were completed using GraphPad Prism to determine statistical significance in demographic variance between the old and new Registry as well between total CHOP and CHOP immunology visits.

## Data Availability

All data produced in the present study are available upon reasonable request to the authors

## Data availability

All USIDNET data is freely available upon approval by the Steering Committee. A standardized query form is on the USIDNET website.

## Acknowledgements

The authors acknowledge all the patients who have taught us over the years.

